# Evaluating prospective study registration and result reporting of trials conducted in Canada from 2009-2019

**DOI:** 10.1101/2022.09.01.22279512

**Authors:** Mohsen Alayche, Kelly D. Cobey, Jeremy Y. Ng, Clare L. Ardern, Karim M. Khan, An-Wen Chan, Ryan Chow, Mouayad Masalkhi, Ana Patricia Ayala, Sanam Ebrahimzadeh, Jason Ghossein, Ibrahim Alayche, Jessie V. Willis, David Moher

**Affiliations:** Centre for Journalology, Ottawa Methods Centre, Ottawa Hospital Research Institute, Ottawa, Canada; Faculty of Medicine, University of Ottawa, Ottawa, Canada; University of Ottawa Heart Institute, Ottawa, Canada; School of Epidemiology and Public Health, Faculty of Medicine, University of Ottawa, Ottawa, Canada; Sport and Exercise Medicine Research Centre, La Trobe University, Melbourne, Australia; Department of Family Practice, University of British Columbia, Vancouver, Canada; Department of Family Practice and School of Kinesiology, University of British Columbia, Vancouver, Canada; Department of Medicine, Women’s College Research Institute, Toronto, Canada; Institute of Health Policy, Management and Evaluation, University of Toronto, Toronto, Canada; School of Medicine, University College Dublin, Dublin, Ireland; Gerstein Science Information Centre, University of Toronto, Toronto, Canada

**Keywords:** Reporting, Trial registration, Trials, Transparency, Reporting best practices, Canadian clinical trials, Publication bias, Clinical trials, Randomized controlled trial

## Abstract

**Background:** Adherence to study registration and reporting best practices are vital to foster evidence-based medicine. Poor adherence to these standards in clinical trials conducted in Canada would be detrimental to patients, researchers, and the public alike.

**Methods:** All registered clinical trials on ClinicalTrials.gov conducted in Canada as of 2009 and completed by 2019 were identified. A cross-sectional analysis of those trials assessed prospective registration, subsequent result reporting in the registry, and subsequent publication of study findings. The lead sponsor, phase of study, clinical trial site location, total patient enrollment, number of arms, type of masking, type of allocation, year of completion, and patient demographics were examined as potential effect modifiers to these best practices.

**Results:** A total of 6,720 trials met the inclusion criteria. From 2009-2019, 59% (n=3,967) of them were registered prospectively and 39% (n=2,642) reported their results in the registry. Of the trials registered between 2009-2014, 55% (n=1,482) were subsequently published in an academic journal. Of the 3,763 trials conducted exclusively in Canada, 3% (n=123) met all 3 criteria of: prospective registration, reporting in the registry, and publishing findings. In contrast, of the remaining 2,957 trials with both Canadian and international sites, 41% (n=1,238) had an overall compliance to these three criteria. Overall, the odds of having adherence to all three practices concurrently in Canadian trials decreases by 95% when compared to international trials (OR = 0.05; 95CI: 0.04 – 0.06).

**Conclusion:** Canadian clinical trials substantially lacked adherence to study registration and reporting best practices. Knowledge of this widespread non-compliance should motivate stakeholders in the Canadian clinical trials ecosystem to address and continue to monitor this problem. The data presented provides a baseline against which to compare any improvement in the registration and reporting of clinical trials in Canada.

## Background

Publication bias is “the tendency on the part of investigators, reviewers, and editors to submit or accept manuscripts for publication based on the direction or strength of the study findings” (1). Studies with statistically significant or positive results are more likely to be published than those with statistically nonsignificant or negative results—this means the dissemination of research findings is a biased process (2–5). Similarly, studies with positive results were much more likely to be published in a shorter time than studies with indefinite conclusions (3). Publication bias threatens the practice of evidence-based medicine, the validity of meta-analyses, and the reproducibility of a study (6). Under-reporting due to publication bias exaggerates the benefits of treatments and underestimates their harms (7). Ultimately, this is detrimental to the healthcare system, as it wastes resources and puts patients at risk (8,9). Between 1999 and 2007, fewer than half of all trials registered and completed on ClinicalTrials.gov were published (10). Between 2010 and 2012, fewer than half of all the registered trials for rare diseases were published within four years of their completion (11).

In 2008, the World Medical Association’s (WMA) Declaration of Helsinki stated that “every clinical trial must be registered in a publicly accessible database before recruitment of the first subject” (12). The declaration also states that all studies should be published, regardless of the statistical significance of their outcome. Nonetheless, publication bias remains prevalent globally. For instance, in all 36 German university medical centers, between 2009 and 2013, only 39% of clinical trials were published within 2 years of their completion (13). In 2015, the World Health Organization (WHO) published a Statement on Public Disclosure of Clinical Trial Results. It states that the main findings of clinical trials are to be published at the latest 24 months after the study completion and that “the key outcomes are to be made publicly available within 12 months of study completion by posting to the results section of the primary clinical trial registry” (14). This WHO statement serves as a global guideline to reduce publication bias and improve overall evidence-based medical decision making.

In 2020, the Canadian Institutes of Health Research (CIHR) asserted that as of 2021, it would implement new policies to ensure full adherence to the WHO Joint Statement requirements (15). Similarly, the Canadian Government recommends adherence to the Tri-Council Policy Statement: Ethical Conduct for Research Involving Humans (TCPS 2), which states that all clinical trials shall be registered before recruitment of the first trial participant, and that researchers shall promptly update the study registry with the location of the findings (16). The Canadian Government also encourages sponsors to register clinical trials in a publicly accessible registry such as ClinicalTrials.gov (17). Despite these recommendations, explicit guidance for trial reporting is lacking (18). Even in jurisdictions where the legal frameworks have been established around trials registration and reporting (e.g., Food and Drug Administration in the United States), low adherence persists (19).

## Objectives

This research aimed to evaluate the adherence of clinical trials conducted in Canada to study registration and reporting best practices. Specifically, among interventional trials registered on clinicaltrials.gov and completed between 2009 and 2019, we aimed to evaluate: (1) the proportion of trials that were prospectively registered on clinicaltrials.gov; (2) the proportion of trials with study results reported on clinicaltrials.gov; (3) the proportion of trials with publication of findings; and (4) characteristics related to clinical trials that may predict prospective registration, reporting of study results, and publication of findings.

## Methods

### Open science statement

Our study protocol was registered on the Open Science Framework (OSF – Link here) prior to data analysis. The data acquired and analyzed in this study are publicly available; therefore, research ethics board approval was not required. We have made our Excel document with all the data, publicly available (link here). We report this cross-sectional study using the STROBE guideline (20).

### Sampling

We obtained our cohort of interventional clinical trials registered in clinicaltrials.gov with a completion date from 2009-19 and with at least one clinical site based in Canada. We chose clinicaltrials.gov because it is the largest, and most comprehensive online registry in the world, currently containing over 423,000 study records from 221 countries (21). The definition of specific terminology on clinicaltrials.gov is outlined in Table 1. Data were downloaded from the registry on October 5^th^ 2021. Microsoft Excel was used to complete the data analysis. To obtain the cohort of Canadian trials, we applied the following filters: A) Interventional Studies (Clinical Trials), B) Completed Studies, C) Canadian Studies D) “Start Date”: 01/01/2009 and E) “Primary Completion Date”: 12/31/2019.

**Table 1.** Definition of certain terms as per clinicaltrials.gov

| Term | Definition |
| --- | --- |
| “Primary Completion Date” | Date on which the last participant in a clinical study was examined or received an intervention to collect final data for the primary outcome measure. |
| “Start Date” | Date on which the first participant was enrolled in a clinical study |
| “Canadian Studies” | All trials with at least one clinical trial site located in Canada, irrespective of the primary investigator’s country of origin |

The time period between 2009 and 2019 was selected because the WMA’s Helsinki statement (published in 2008) gave all researchers an opportunity to become familiar with best practices for study registration. It often takes more than 5 years from inception to publish a completed clinical trial (3). Therefore, when evaluating studies for publication in an academic journal, we excluded any study with a primary completion date after 12/31/2014 so that all researchers had at least 5 years to publish their results.

### Outcomes

For all registered and completed clinical trials conducted in Canada between 2009 and 2019, we counted the proportion that: (1) prospectively registered their study before the recruitment of their first participant; (2) reported their results in the registry and (3) published study findings.

The first two outcomes were measured directly by analyzing the data downloaded from clinicaltrials.gov. Prospective registration and reporting trial results in clinicaltrials.gov was taken directly from the registry. Assessing whether the trial findings were published in an academic journal was more complicated than the first two outcomes. Notification of publishing the trial finding in a journal requires additional effort and some degree of interpretation.

To measure the third outcome, we added the following search criteria “(NOT NOTEXT) [CITATIONS]” in the “Other terms” box in clinicaltrials.gov to identify clinical trials with publications in an academic journal. Those publications were either automatically indexed by clinicaltrials.gov, or manually added by the sponsor or investigator. As per the clinicaltrials.gov website, citations are automatically identified by clinicaltrials.gov based on their NCT number and are subsequently indexed in the database. Clinicaltrials.gov does not expand any further on how these studies are automatically identified and indexed (22).

### Trial characteristics

A total of ten study characteristics were recorded for each trial including: lead sponsor (e.g., industry); phase of clinical trial (e.g., phase 3); total number of participants e.g., <100); biological sex of participants (e.g., male); primary completion date (e.g., 2019); clinical trial site location (e.g., Canada); number of arms (e.g., two-arm trial); type of masking (e.g., double blind); intervention model (e.g., parallel assignment); type of allocation (e.g., randomized).

## Data Analysis

We calculated the proportions of trials that were prospectively registered, had their results reported in the registry, and subsequently had their findings published.

We analyzed whether the results were modified by these 10 variables of interest as outlined in the study characteristics above using univariable logistic regressions (odds ratios with 95% confidence intervals). This analysis enables us to identify which characteristics have the strongest impact on best practice adherence. The outcomes of this analysis were reported as a prevalence in percentages.

### Quality Assurance

To verify the clinicaltrials.gov records of publication status, we randomly selected a 10% sample of studies for manual verification. The 10% sample was randomly picked for quality assurance via the Excel “RAND” function. Each study in the sample was manually searched in a 3-step process by four members of the research team: (1) the clinical trial identifier (NCT ID) was entered on clinicaltrials.gov to verify the publication status posted on the website; (2) the NCT ID was then searched on PubMed to verify the publication status; and (3) the NCT ID was finally searched on Google Scholar to verify the publication status. We confirmed the lack of publication of a study if both the PubMed and Google Scholar search yielded no results. We confirmed the successful publication of a study if either PubMed or Google Scholar yielded results.

## Results

A total of 6,790 clinical trials conducted in Canada were identified. Of those clinical trials, we excluded 70 of them that were submitted to clinicaltrials.gov with one or more incorrect entries and were therefore incompatible with our software (Microsoft Excel). For example, among those 70 studies, a few had submitted a “alphabetic” entry despite a strictly “numerical” requirement. Our software could not correct those mistakes and therefore those studies were excluded. Therefore, a total of 6,720 (99%) studies were included in our analysis.

### Demographic data

The demographic data of the collected sample are highlighted in Table 2. The median year of trial primary completion was 2015. Of the 6720 included clinical trials, 1% (n=65) had a primary completion date in 2009; 12% (n=819) had a primary completion date in 2019. A total of 38% (n=2,581) of identified trials did not indicate the phase of their study. 59% (n=3,967) of trials were prospectively registered, while the remaining trials were registered after recruitment started. Moreover, 39% (n=2,642) of all trials made their results available in the registry.

**Table 2.** Trial characteristics

|  |  |  |
| --- | --- | --- |
| **Total** | **6,720** | **100.0%** |
| **Time of Registration** |  |  |
| Before trial start | 3967 | 59% |
| After trial start | 2753 | 41% |
| **Results Reported** |  |  |
| Results reported in the registry | 2642 | 39% |
| Results not reported in the registry | 4078 | 61% |
| **Publication of findings** |  |  |
| Published | 3724 | 55% |
| Not published | 2996 | 45% |
| **Lead Sponsor** |  |  |
| Industry<sup>±</sup> | 3245 | 48% |
| Academia<sup>*</sup> | 3475 | 52% |
| **Phase** |  |  |
| Early I | 36 | 1% |
| I | 490 | 7% |
| I-II | 181 | 3% |
| II | 1256 | 19% |
| II-III | 98 | 1% |
| III | 1587 | 24% |
| IV | 491 | 7% |
| Not reported | 2581 | 38% |
| **Number of participants** |  |  |
| 1-99 | 3496 | 52% |
| 100-500 | 2182 | 32% |
| >500 | 1041 | 15% |
| Not reported | 1 | 0% |
| **Clinical trial site location** |  |  |
| Canada only | 3763 | 56% |
| International<sup>§</sup> | 2957 | 44% |
| **Primary completion date** |  |  |
| 2009 | 65 | 1% |
| 2010 | 216 | 3% |
| 2011 | 399 | 6% |
| 2012 | 554 | 8% |
| 2013 | 681 | 10% |
| 2014 | 736 | 11% |
| 2015 | 787 | 12% |
| 2016 | 781 | 12% |
| 2017 | 848 | 13% |
| 2018 | 834 | 12% |
| 2019 | 819 | 12% |

<sup>±</sup> The industry category includes pharmaceutical and device companies and NIH studies.
<sup>\*</sup> The academia category includes universities, individuals, and community-based organization.
<sup>§</sup> Includes all international clinical trials that have at least one Canadian site.

### Primary Completion Date

Outlined in Table 3 are all the primary outcomes we measured based on the year of primary completion. From 2009 to 2019, there was an increasing number of studies reaching primary completion. There was an increasing trend from 2009-2019 in the prevalence of prospective registration: 35% in 2009 and 73% in 2019. However, there is no trend observed in the reporting of results across the years: 34% in 2009 and 32% in 2019. Overall, there was a slight upward trajectory in the adherence to all three practices, mostly explained by the increase in prospective registration and publication of findings.

**Table 3.** Adherence of Canadian trials to study registration and reporting best practices based on the year of primary completion.

| **Year of completion** | **Number of studies** | **Prospective registration** | **Results reported** | **Findings published<sup>\*</sup>** | **All three practices<sup>\*</sup>** |
| --- | --- | --- | --- | --- | --- |
| 2009 | 65 | 23 (35.38%) | 22 (33.85%) | 24 (36.92%) | 3 (4.62%) |
| 2010 | 216 | 92 (42.59%) | 82 (37.96%) | 120 (55.56%) | 30 (13.89%) |
| 2011 | 399 | 174 (43.61%) | 161 (40.35%) | 210 (52.63%) | 74 (18.55%) |
| 2012 | 554 | 276 (49.82%) | 215 (38.81%) | 304 (54.87%) | 101 (18.23%) |
| 2013 | 681 | 354 (51.98%) | 305 (44.79%) | 386 (56.68%) | 145 (21.29%) |
| 2014 | 736 | 413 (56.11%) | 302 (41.03%) | 438 (59.51%) | 174 (23.64%) |
| 2015 | 787 | 453 (57.56%) | 341 (43.33%) | - | - |
| 2016 | 781 | 472 (60.44%) | 308 (39.44%) | - | - |
| 2017 | 848 | 542 (63.92%) | 347 (40.92%) | - | - |
| 2018 | 834 | 573 (68.71%) | 300 (35.97%) | - | - |
| 2019 | 819 | 595 (72.65%) | 259 (31.62%) | - | - |
| Total | 6720 | 3967 (59.03%) | 2642 (39.32%) | 1482 (55.9%) | 527 (19.88%) |

<sup>\*</sup>Years 2015-2019 were excluded when measuring the “published” variable.

### Lead sponsor, phase of study, total enrollment and countries implicated

Table 4 outlines all the primary outcomes based on lead sponsor, total patient enrollment, phase of study, and clinical trial site location. Trials with an “Industry” lead sponsor had higher rates of prospective registration, reporting of results, and publication of study findings than trials with an “Academia” lead sponsor. Overall clinical trials led by the “Industry” had a 36% (n=1,182) adherence to all three best practices while clinical trials led by “Academia” had an adherence of only 5% (n=179). A univariable analysis determined that the odds of having prospective registration with an “Academia” lead sponsor decreases by 56% when compared to “Industry” (OR = 0.44; 95CI: 0.40 – 0.49). Moreover, the odds of result reporting are 93% lower (OR = 0.07; 95CI: 0.06 – 0.08), and publication of findings is 13% lower (OR = 0.87; 95CI: 0.79 – 0.96). Overall, adherence to all three practices concurrently is 90% lower in “Academia” than in “Industry” (OR = 0.1; 95CI: 0.09 – 0.12).

**Table 4.** Adherence of Canadian trials to study registration and reporting best practices based on the lead sponsor, total enrollment, phase of study, and countries implicated

| Variable of Interest | Number of studies | Prospective registration | Results reported | Findings published | All three practices |
| --- | --- | --- | --- | --- | --- |
| Total | 6720 | 3967 (59.03%) | 2642 (39.32%) | 3724 (55.42%) | 1361 (20.25%) |
| **Lead sponsor** |  |  |  |  |  |
| Industry<sup>±</sup> | 3245 | 2235 (68.88%) | 2217 (68.32%) | 1849 (56.98%) | 1182 (36.43%) |
| Academia* | 3475 | 1732 (49.84%) | 425 (12.23%) | 1875 (53.96%) | 179 (5.15%) |
| **Total patient enrollment** |  |  |  |  |  |
| 1-99 | 3496 | 1813 (51.86%) | 762 (21.8%) | 1538 (43.99%) | 274 (7.84%) |
| 100-500 | 2182 | 1402 (64.25%) | 1101 (50.46%) | 1355 (62.1%) | 585 (26.81%) |
| >500 | 1041 | 752 (72.24%) | 779 (74.83%) | 831 (79.83%) | 502 (48.22%) |
| Not given | 1 | 0 (0%) | 0 (0%) | 0 (0%) | 0 (0%) |
| **Phase of study** |  |  |  |  |  |
| Early I | 36 | 14 (38.89%) | 3 (8.33%) | 14 (38.89%) | 0 (0%) |
| I | 490 | 262 (53.47%) | 73 (14.9%) | 145 (29.59%) | 19 (3.88%) |
| I/II | 181 | 97 (53.59%) | 51 (28.18%) | 79 (43.65%) | 18 (9.94%) |
| II | 1256 | 863 (68.71%) | 679 (54.06%) | 634 (50.48%) | 322 (25.64%) |
| II/III | 98 | 60 (61.22%) | 34 (34.69%) | 61 (62.24%) | 16 (16.33%) |
| III | 1587 | 1172 (73.85%) | 1251 (78.83%) | 1192 (75.11%) | 777 (48.96%) |
| IV | 491 | 295 (60.08%) | 197 (40.12%) | 285 (58.04%) | 106 (21.59%) |
| Not given | 2581 | 1204 (46.65%) | 354 (13.72%) | 1314 (50.91%) | 103 (3.99%) |
| **Clinical trial site location** |  |  |  |  |  |
| Canada only | 3763 | 1774 (47.14%) | 435 (11.56%) | 1800 (47.83%) | 123 (3.27%) |
| International<sup>§</sup> | 2957 | 2193 (74.16%) | 2207 (74.64%) | 1924 (65.07%) | 1238 (41.87%) |

<sup>±</sup> The industry category includes pharmaceutical companies, device companies, and NIH studies.
\* The academia category includes universities, individuals, and community-based organization.
<sup>§</sup> Includes all international clinical trials that have at least one Canadian clinical site.

There was a higher adherence to study registration and reporting best practices based on the size of the clinical trial. Of the clinical trials with over 500 participants, 48% (n=502) adhered to all three practices. Clinical trials with less than 100 participants had an overall adherence of only 8% (n=274). A univariable analysis determined that the odds of prospective registration of trials with <100 participants decreases by 59% when compared to trials with >500 participants (OR = 0.41; 95CI: 0.36 – 0.48). Moreover, the odds of result reporting is 91% lower (OR = 0.09; 95CI: 0.08 – 0.11), and publication of findings is 80% lower (OR = 0.20; 95CI: 0.17 – 0.23). Overall, adherence to all three practices concurrently is 91% lower in trials with <100 participants than in trials with >500 participants (OR = 0.09; 95CI: 0.08 – 0.11).

Phase 3 trials had the highest performance of prospective registration, result reporting, and publication of findings in comparison to any other phase. Phase 3 studies have an adherence of 49% (n=777) to all three practices, as opposed to phase 1 studies with an adherence of 4% (n=19). A univariable analysis determined that the odds of prospective registration with a Phase 1 trial decreases by 59% when compared to Phase 3 trial (OR = 0.41; 95CI: 0.33 – 0.50). Moreover, the odds of result reporting is 95% lower (OR = 0.05; 95CI: 0.04 – 0.06), and publication of findings is 86% lower (OR = 0.14; 95CI: 0.11 – 0.17). Overall, adherence to all three practices concurrently is 96% lower in Phase 1 trials than in Phase 3 trials (OR = 0.04; 95CI: 0.03 – 0.07).

International clinical trials had a higher rate of prospective registration 74% (n=2,193), result reporting 75% (n=2,207) and publication of findings 65% (n=1,924) than trials conducted with exclusively Canadian sites. Overall, International trials had a 42% (n=1,238) adherence to all three practices and “Canadian Only” trials had an adherence of 3% (n=123) to all three practices. A univariable analysis determined that the odds of having prospective registration by Canadian trials decreases by 69% when compared to international trial (OR = 0.31; 95CI: 0.28 – 0.35). Moreover, the odds of result reporting is 96% lower (OR = 0.04; 95CI: 0.04 – 0.05), and publication of findings is 51% lower (OR = 0.49; 95CI: 0.45 – 0.54). Overall, adherence to all three practices concurrently is 95% lower in Canadian trials than in international trials (OR = 0.05; 95CI: 0.04 – 0.06).

The remaining results of the variables of interest are in the appendix. All the data analyzed, the statistical analyses, and the results pertaining to the top Canadian institutions can be accessed directly via this link here.

### Quality Assurance Results

A random 10.0% sample (n=672) was selected for manual verification of the publication status. As per the downloaded data from clinicaltrials.gov, 55% (n=371) of those trials had results published in a medical journal. The quality assurance determined that 69% (n=463) of those trials were in fact published.

Clinicaltrials.gov underestimated the true prevalence of published clinical trials because: (1) some trials were published without an NCT ID; (2) some trials were published after we downloaded the data; (3) some trials were published in the form of an Abstract/Thesis/Poster/PhD; and (4) Clinicaltrials.gov was not able to automatically index some publications in certain journals.

## Discussion

Of the almost 7000 RCTs in our sample, less than two-thirds were prospectively registered; less than half made their results available; and less than two-thirds published their findings. Less than a quarter of the RCTs completed all three best practices. Trials conducted with exclusively Canadian sites were substantially less compliant to these practices than trials with both Canadian and international sites. Importantly, the trials we describe were not small (23); nearly a third of the trials included between 100 and 500 participants and 15% (n=1041) of them included over 500 participants.

Of all the variables included in the logistic regression, four were highly associated with study registration and reporting best practices. They were: (1) clinical trials led by “Industry” (pharmaceutical companies), (2) phase 3 clinical trials, (3) trials with over 500 participants and finally (4) clinical trials conducted by a multinational team. The four variables that were negatively associated with study registration and reporting best practices were: (1) clinical trials led by “Academia” (universities), (2) phase 1 clinical trials, (3) trials with fewer than 100 participants and (4) trials conducted with exclusively Canadian sites.

Some readers will view these results as another example of egregious waste in biomedicine with little improvement since the 2014 landmark Lancet series on research waste (24). Patients, who are critical to the success of clinical trials, are likely to be disappointed with these results; their contributions are not being honored. For clinical practice guideline developers, these results indicate that evidence is missing regarding the totality of knowledge about an intervention. For healthcare funders, these results indicate a bad return on investment. If grantees use scarce resources, often taxpayer dollars, and do not prospectively register their trials and/or make their results available in registries or publish their results, everyone loses. Finally, academic institutions may risk institutional reputation when their faculty members fail to meet minimum national/global standards (WHO; CIHR).

These results are not unique to Canada. Similar results have been reported elsewhere (13,25–27). The overall lack of prospective registration and reporting clinical trial results may reflect a lack of knowledge on the topic on the part of clinical trials teams and/or their academic institutions. Prospective registration and reporting results are part of a larger ecosystem of open science, which includes transparency. Compared to some other parts of the world, Canada has been slow to publicly embrace the practices of open science (28).

The lack of prospective registration may reflect that clinical trial principal investigators are leaving these responsibilities to other team members. Alternatively, funders may not have strong enough adherence monitoring in place. With the advent of automated digital monitoring and the increasing availability of Application Programming Interfaces (APIs) this should be less of a problem. The European Trials Tracker scheme, developed by the University of Oxford’s Bennett Institute for Applied Data Science is an example of existing monitoring on a large scale (https://www.bennett.ox.ac.uk/) (29). Funders and academic institutions could use digital dashboards to monitor adherence to clinical trial policies and identify training needs (30).

In the last few years, several initiatives have proposed moving away from traditional metrics of the number of publications (the irony of our results—counting publications—is not lost on us) towards a broader set of best practices that reflect an institutional commitment to research integrity when assessing researchers for hiring, promotion and tenure (31–33). Current researcher assessment could be augmented by tracking whether faculty members have prospectively registered their trials (and other studies), made their results available on a trial register, and published them (preferably in an open access journal).

It is time for trial funders and academic institutions to collaborate to address the overall lack of adherence of Canadian trials to registration and reporting mandates. In the Canadian context, there is now a requirement for equity, diversity, and inclusion (EDI) training when applying for grant funding for the CIHR. CIHR has had success in requiring principal investigators to take sex and gender training before they can submit a grant application (34). Something similar could be introduced for clinical trial registration and results reporting. The training can be required for all faculty and research staff involved in conducting trials. This would portray the funders and academic institutions’ commitment to improve this situation. they could also collectively commit to evaluating such an educational intervention by conducting a stepped wedge and/or cluster trial across universities to ensure the training was having the desired effect.

From the earliest attempts to introduce clinical trial registration in the 1980s there was a strong belief that it might reduce publication bias and provide a more accurate picture of the estimated benefits and harms effect of interventions. Our results, and others (13,35), suggest that publication bias is still a substantive problem despite prevalent views that clinical trials are heavily regulated. Indeed, although regulation via policy exists, if we fail to audit adherence, the policy goals will not be adhered to.

CIHR recently updated their policy guide (see box). The ‘stick’ in this updated guidance is likely to be the lack of future funding for principal investigators unless their current trials are registered, and the results made available. Importantly, the agency will monitor adherence annually “by asking impacted researchers to provide clinical trial registration identifiers, and links to summary results and open access publications.”

“The following new requirements apply to all clinical trial grants funded on or after January 1, 2022:

- Public disclosure of results must be done within a mandated time frame:
  - publications describing clinical trial results must be open access from the date of publication;
  - summary results must be publicly available within 12 months from the last visit of the last participant (for collection of data on the primary outcome); and
- All study publications must include the registration number/Trial ID (to be specified in the article summary/abstract).

Nominated Principal Investigators receiving CIHR grant funds for clinical trial research after January 1, 2022, must comply with the above requirements in order to remain eligible for any new CIHR funding.”

## Limitations

We relied on the data as reported in clinicaltrials.gov. This is the most widely used registration platform (36) accounting for the vast majority of all clinical trial registrations. To be counted in our analysis, we only tracked forward the trials that were marked as ‘completed’ on clinicaltrials.gov; this means we missed records that were registered and then never updated to be marked as complete despite these trials having ended. The results are likely worse than we report here. Moreover, we only tracked studies that were registered on clinicaltrials.gov in the first place. Had we also analyzed publications that were not registered in clinicaltrials.gov, adherence may have been lower. Furthermore, we did not analyze the length of time it took between the study completion date and the publication date. In other words, some studies may have posted their results and published their findings a decade after the study completion despite the recommendation to do so within two years (14). Despite all of this, less than one quarter of the sample adhered to all three best practices. Some may argue that trials with non-statistically significant results take longer to publish, however, recent review found no difference in time to publication by the statistical significance of the trial (37).

Another limitation is that we relied on clinicaltrials.gov regarding the publication of study findings. Ideally, we would have completed the quality assurance to our entire sample, however due to a resource constraint, we limited the quality assurance to 10% of our sample. As demonstrated by our quality assurance, clinicaltrials.gov has underestimated the true prevalence of publications by roughly 14%. Clinicaltrials.gov reported that 55% (n=371) of trials were published, but our quality assurance determined the true number to be 69% (n=463). Some publications were missed by clinicaltrials.gov for the following reasons: publication does not include NCT IDs, publication date was after we collected the data, publications were in the form of an Abstract/Thesis/Poster, and finally some publications were in journals not accessible by clinicaltrials.gov. Overall, this underestimation of publication status supports our findings.

In summary, our analysis of nearly 7000 Canadian trials registered on clinicaltrials.gov indicates that there is substantial room for improvement in ensuring they are prospectively registered at inception (prior to the first person be randomized), the results are reported in a publicly accessible registry, and that completed trials are published, preferable in an open access platform/journal. The consequences of not monitoring adherence to these activities is profound and wasteful. The international AllTrials initiative, signed by more than 700 organizations, has brought attention for public support for these policies (38). Several Canadian-based foundations have signed this declaration (e.g., Canadian Cancer Research Alliance, Canadian Agency for Drugs and Technologies in Health, Canadian Medical Association, Canadian HIV Trials Network). Other stakeholders like Health Canada and funders of academic trials like the federal Tri Agency ought to also commit to the AllTrials initiative and its broader principles. Canada should join the global movement to address research waste due to incompliant registration and reporting of trials and seek to lead in identifying resolutions.

## Data Availability

All data referred to in the manuscript are available online at https://osf.io/kb4nf/

## Funding

This work was supported by the Summer Studentship Program from the University of Ottawa Faculty of Medicine.

## Conflicts of interest

The authors declare no conflicts of interest.

## Author contributions

Conceptualization: DM, KDC, MA

Study Design: DM, KDC

Funding acquisition: MA

Project administration: MA

Supervision: DM, KDC

Writing – original draft: MA

Writing – review & editing: All authors

## Appendix

### Variable of Interest: Biological Sex of Participants

**Table 5.** Adherence of Canadian trials to study registration and reporting best practices based on biological sex of participants.

| **Biological Sex of Participants** | **Total Number of Studies** | **Prospective Registration** | **%** | **Results Reported** | **%** | **Published** | **%** | **All 3 Practices** | **%** |
| --- | --- | --- | --- | --- | --- | --- | --- | --- | --- |
| All | 5932 | 3572 | 60.22% | 2403 | 40.51% | 3319 | 55.95% | 1265 | 21.33% |
| Female | 493 | 241 | 48.88% | 143 | 29.01% | 255 | 51.72% | 49 | 9.94% |
| Male | 295 | 154 | 52.20% | 96 | 32.54% | 150 | 50.85% | 47 | 15.93% |
| **Total** | **6720** | **3967** | **59.03%** | **2642** | **39.32%** | **3724** | **55.42%** | **1361** | **20.25%** |

### Variable of Interest: Type of Masking

**Table 6.** Adherence of Canadian trials to study registration and reporting best practices based on type of masking.

| **Masking** | **Total Number of Studies** | **Prospective Registration** | **%** | **Results Reported** | **%** | **Published** | **%** | **All 3 Practices** | **%** |
| --- | --- | --- | --- | --- | --- | --- | --- | --- | --- |
| None | 2950 | 1706 | 57.83% | 1040 | 35.25% | 1450 | 49.15% | 469 | 15.90% |
| Single | 888 | 421 | 47.41% | 142 | 15.99% | 509 | 57.32% | 60 | 6.76% |
| Double | 1019 | 650 | 63.79% | 506 | 49.66% | 603 | 59.18% | 274 | 26.89% |
| Triple | 624 | 377 | 60.42% | 293 | 46.96% | 356 | 57.05% | 160 | 25.64% |
| Quadruple | 1224 | 809 | 66.09% | 659 | 53.84% | 797 | 65.11% | 397 | 32.43% |
| Not reported | 15 | 4 | 26.67% | 2 | 13.33% | 9 | 60.00% | 1 | 6.67% |
| **Total** | **6720** | **3967** | **59.03%** | **2642** | **39.32%** | **3724** | **55.42%** | **1361** | **20.25%** |

### Variable of Interest: Intervention Design

**Table 7.**
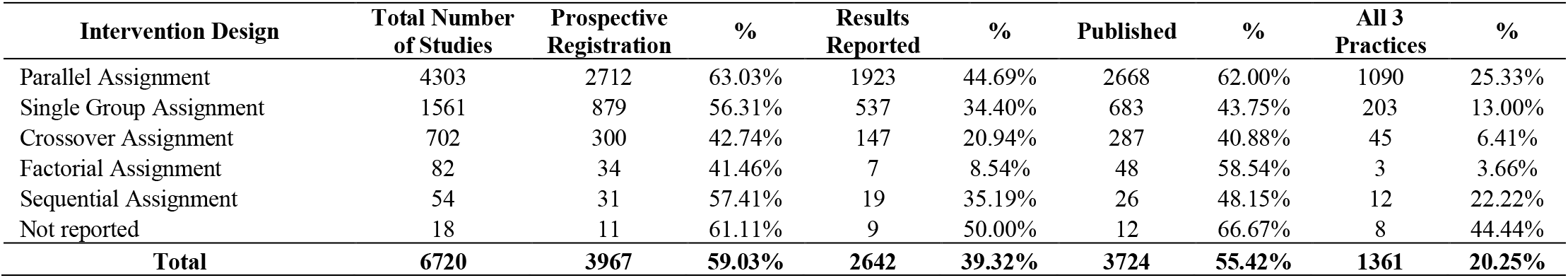
Adherence of Canadian trials to study registration and reporting best practices based on intervention.

### Variable of Interest: Allocation

**Table 8.** Adherence of Canadian trials to study registration and reporting best practices based on allocation.

| **Allocation** | **Total Number of Studies** | **Prospective Registration** | **%** | **Results Reported** | **%** | **Published** | **%** | **All 3 Practices** | **%** |
| --- | --- | --- | --- | --- | --- | --- | --- | --- | --- |
| Randomized | 4965 | 2954 | 59.50% | 2002 | 40.32% | 2955 | 59.52% | 1106 | 22.28% |
| Non-Randomized | 498 | 304 | 61.04% | 197 | 39.56% | 226 | 45.38% | 86 | 17.27% |
| N/A | 1235 | 699 | 56.60% | 437 | 35.38% | 533 | 43.16% | 166 | 13.44% |
| Not reported | 22 | 10 | 45.45% | 6 | 27.27% | 10 | 45.45% | 3 | 13.64% |
| **Total** | **6720** | **3967** | **59.03%** | **2642** | **39.32%** | **3724** | **55.42%** | **1361** | **20.25%** |

### Variable of Interest: Arms

**Table 9.** Adherence of Canadian trials to study registration and reporting best practices based on number of arms.

| Number of Arms | Total Number of Studies | Prospective Registration | % | Results Reported | % | Published | % | All 3 Practices | % |
| --- | --- | --- | --- | --- | --- | --- | --- | --- | --- |
| 1 | 1302 | 733 | 56.30% | 467 | 35.87% | 563 | 43.24% | 180 | 13.82% |
| 2 | 3555 | 2127 | 59.83% | 1279 | 35.98% | 2081 | 58.54% | 696 | 19.58% |
| 3 | 910 | 551 | 60.55% | 446 | 49.01% | 542 | 59.56% | 246 | 27.03% |
| 4 | 473 | 274 | 57.93% | 222 | 46.93% | 277 | 58.56% | 118 | 24.95% |
| Not reported | 60 | 27 | 45.00% | 1 | 1.67% | 23 | 38.33% | 0 | 0.00% |
| **Total** | **6720** | **3967** | **59.03%** | **2642** | **39.32%** | **3724** | **55.42%** | **1361** | **20.25%** |

\*A total of 480 clinical trials in our sample had a “number of arms” reported between 5 and 24. Those trials were excluded from this table. The “Total” row however reflects the entire sample (n=6720).

## Notes

### Competing Interest Statement

The authors have declared no competing interest.

### Clinical Protocols

https://osf.io/kb4nf/

### Funding Statement

This work was funded by the Summer Studentship Program from the University of Ottawa, Faculty of Medicine.

